# Expanding COVID-19 symptom screening to retail, restaurants, and schools by preserving privacy using relaxed digital signatures

**DOI:** 10.1101/2020.08.06.20169839

**Authors:** Brandon Jew, Alexis Korb, Paul Lou, Jeffrey N. Chiang, Ulzee An, Amit Sahai, Eran Halperin, Eleazar Eskin

## Abstract

Symptom screening is a widely deployed strategy to mitigate the COVID-19 pandemic and many public health authorities are mandating its use by employers for all employees in the workplace. While symptom screening has the benefit of reducing the number of infected individuals in the workplace, it raises some inherently difficult privacy issues as a traditional approach requires the employer to collect symptom data from each employee which is essentially medical information. In this paper, we describe a system to implement Cryptographic Anonymous Symptom Screening (CASS) which allows for individuals to perform COVID symptom screening anonymously while avoiding the privacy issues of traditional approaches. In the system, individuals report their symptoms without any identifying information and are issued a completion certificate. This certificate contains a cryptographic code which certifies that the certificate was obtained from the screener after reporting no symptoms. The codes can be verified using a cryptographic algorithm which is publicly available. A standard cryptography approach to implement such a system would be to use digital signatures. Unfortunately, standard digital signatures have some limitations for this application in that the signatures are often hundreds of characters long and if the signature contains the name of the individual, then there is also a risk of compromising privacy. In our approach, we develop and utilize a relaxed digital signature scheme to provide 16 character long codes and handle names using equivalence classes which helps preserve privacy. Both of these extensions technically compromise the security but in a way that is negligible for this application. Our system can either serve the function of standard symptom screening system approaches for employees, but can also extend symptom screening to non-employees such as visitors or customers. In this case, the system can be utilized in retail, restaurants and schools to ensure that everyone in the physical space, including employees, customers, visitors and students have performed symptom screening.

## Introduction

As we start the process of reopening the economy and our society, critical infrastructure is being put into place to help manage the pandemic to suppress infection rates. This includes the widespread availability of testing, infrastructure for contact tracing, and symptom screening. Many public health authorities are mandating employers to implement workplace symptom screening. Symptom data has been widely collected to measure the state of the pandemic through surveys (Menni et al. 2020; Segal et al. 2020; Timmers et al. 2020; Drew et al., n.d.) and analysis of this data combined with studies that collect symptoms in conjunction with diagnostic testing estimate that 60% of COVID-19 positive individuals have symptoms characteristic of infection (Nishiura et al. 2020; Mizumoto et al. 2020; Day 2020).

These efforts have reinforced the need for systematic screening based on symptoms which can identify over half of COVID-19 infections. Such screening is used to help self-identify individuals with symptoms to reduce the number of individuals with infection in a workplace. Each day before reporting to a workplace, employees complete an online survey which asks for symptom information. Employees who are symptom free receive a certificate that can be used to gain entry to the workplace. Employees who report symptoms are prevented from coming to the workplace and prioritized for testing. These types of surveys are commonly performed by health care employers for their workers (Zhang et al. 2020; Lan et al. 2020; Tostmann et al. 2020; Yombi et al. 2020). For example, UCLA Health implemented such a system for its health care workers in early April. Other health systems have similar procedures and these efforts have been shown to identify individuals who are infected protecting other workers and patients. UCLA implemented a similar system for its campus in late April.

Symptom screening is also considered a strategy to reduce risk when traveling (Gostic et al. 2020).

There are some inherent logistical, privacy and legal-related challenges to implementing workplace symptom screening (Kitchin 2020). When an employer deploys a symptom screening system, the employer is collecting and storing medical information on the employees. There are also complex legal and privacy issues related to who can access that information and what happens if the information is accessed inappropriately. In addition, employees are not the only individuals in a workplace. Customers, visitors and other individuals may also be carriers of infection, yet not subject to symptom screening. However, how to address logistical, privacy and legal-related challenges for non-employees are potentially even more challenging. Despite these challenges, symptom screening does have a clear positive effect on reducing infection rates.

We propose a technological solution to the privacy issues in performing workplace symptom screening, Cryptographic Anonymous Symptom Screening (CASS). In our solution, participants will use a third party symptom screener anonymously. They provide symptom information to the third party hosted system but not any identifying information. If they are symptom free, they are issued a symptom free completion certificate that provides evidence that they took the screener and reported that they are symptom free and also records a specific hour of the day that the screener was taken. As no identifying information is collected by the screener, the certificate contains no identifying information. We refer to this type of certificate as an anonymous completion certificate. The certificate can then be shared with the employer in order to gain access to the workplace. The certificate contains a type of “digital signature” issued by the screener which certifies that the individual completed the survey and reported no symptoms.

The digital signature can be verified as valid by the employer. Digital signatures are a widely utilized cryptographic technique (Rivest, Shamir, and Adleman 1978) with many applications including public-key infrastructures (PKI) in the form of TLS/SSL. Since it is virtually impossible to forge a valid digital signature, by verifying the signature, an employer can confirm that an individual reported as symptom free to the third party screener.

Unfortunately, standard digital signatures have some limitations for this application in that the signatures are often hundreds of characters long. Such a long signature is not practical to use on a daily symptom free certificate. Some individuals will print their certificates so that the digital signatures will need to be entered manually into a verification system. Other individuals would want their signature embedded in a URL. In our approach, we present a novel relaxed digital signature scheme to provide 16 character long codes. The tradeoff is that while standard digital signatures can encode an exact timestamp, our codes can only encode a specific hour of the day. However, for our application, this tradeoff is completely acceptable since we are interested in knowing whether or not an individual is symptom free in the last 24 hours and do not need the exact time. Our cryptographic algorithm still makes it impossible to guess a valid code, similar to standard digital signatures, which prevents circumventing taking the survey.

Everyone who completes the screener in a specific hour and reports no symptoms will receive the same completion code on their certificates. Even if the employer is able to obtain access to the third party symptom screener (which they can not) they would not be able to distinguish between the many individuals who share the same code. While it is possible for someone to complete the screener and then share their anonymous certificate with someone else to avoid taking the screener, we feel that this is acceptable because this gives confidence to the participants that their privacy is protected. In practice, it is much easier for an individual to fill out the symptom survey than to obtain someone else’s anonymous certificate.

In some environments, the concern about sharing anonymous certificates is unacceptable and they would prefer certificates with identifying information on the certificate. Our system can also issue symptom free completion certificates with an individual’s name or other identifier on the certificate. This is also possible with standard digital signatures as it is standard to include a name in the signature which can be validated. If the name is changed on the certificate then the signature will no longer match and can easily be identified as invalid. Unfortunately, using standard digital signatures then undermines the privacy protection of the system as the third party screener will need to know the name of an individual filling out the survey to create the digital signature. To preserve privacy in our approach, we extend standard digital signatures to use name equivalence classes which are groupings of names. All certificates issued during a specific hour with names within an equivalence class will have the same code. The third party screener does not obtain the actual name of the individual but only the class and if there are many people taking the survey each hour, there will be many individuals in each class so privacy will be preserved. If an individual tries to change the name on a certificate, it will only still be valid if the two names are in the same class, but with the large number of classes, this is unlikely to be practical and has a negligible effect on security.

We note that because CASS preserves privacy, CASS has the potential to have additional applications beyond employee symptom screening. For example, retail stores or restaurants can use such a system for symptom screening their customers. Schools can use such a system for parents to report the symptoms of their children before bringing them to school.

At UCLA, we implemented CASS and have made the system available to the community. In our implementation, the symptom free completion certificates are designed so that they can be displayed on a mobile device and they change color over time. A completion certificate is blue when the symptom survey was taken in the last 24 hours. It turns yellow between 24 hours and one week and turns red after one week.

## CASS System Overview

We describe our system by first contrasting to traditional symptom screening. In traditional symptom screening, there are two relevant entities. We refer to the individual who is screening themselves as the user. We refer to the organization that is requesting that the screening is being performed as “the organization” which is often an employer, but could also be a university or other entity that wants to make their location safer. The organization implements a screening system which is usually a software that they install which collects user symptom information and if a user is symptom free, they are issued certificates that clears them to be able to enter the workplace. The organization maintains the database of user symptom information which creates some legal and privacy challenges.

Our approach, Cryptographic anonymous symptom screening (CASS), allows for symptom screening without compromising privacy which is enabled by extending digital signatures in two ways. In CASS, there is a third relevant entity which is a third party screener that hosts the screening system and a fourth entity which is the verification system. Users report their symptoms to the third party screener and if the user reports that they are symptom free, they receive a symptom free certificate providing evidence that they completed the screener and reported no symptoms. Users can show this to their organization and the organization can verify the authenticity of the certificate using the verification system. In order to preserve privacy, a few conditions must hold. First, the user only shares symptom information with the screener, but not any identifying information. If the user is symptom free, the user receives a symptom free completion certificate which can be shared with the organization. Second, other than obtaining the certificate directly from the user, the organization does not receive any symptom information about the user. Third, the organization can not access the symptom data of the users.

For the system to be effective, the organization needs to have some guarantee that if a user has possession of a symptom free certificate, the user entered symptom free data into the screener and obtained a certificate. We use a novel relaxed digital signature scheme for this application. The certificates issued by the screener contain code which is effectively a digital signature so that they can be verified as authentic by the organization using the verification system. Unlike traditional digital signatures, our codes are only 16 characters long.

We implemented CASS at UCLA and provide both a third party screener and a verification system to support anonymous and named certificates. In our implementation of the system at UCLA, the verification system visualizes the code as a certificate taking advantage of the fact that the code encodes the time the symptom screening was performed. On the certificate, the code is both written and encoded in a QR code. A code can be verified to be authentic using the verification system. Similar to digital signatures, because of the cryptographic algorithm, it is extremely difficult to obtain a valid code without getting it from the system. In our implementation, the color of the certificate shows how recent the screening was performed. Blue signifies within 24 hours, yellow signifies within a week and red is older than one week. On the certificate is also the time that the screening was performed. The certificate is designed to be displayed on a mobile phone but can also be printed out. Using a mobile device, the QR code on a certificate can be scanned to verify that it is valid. Examples of anonymous certificates are shown in Figure 1.

**Figure 1.**
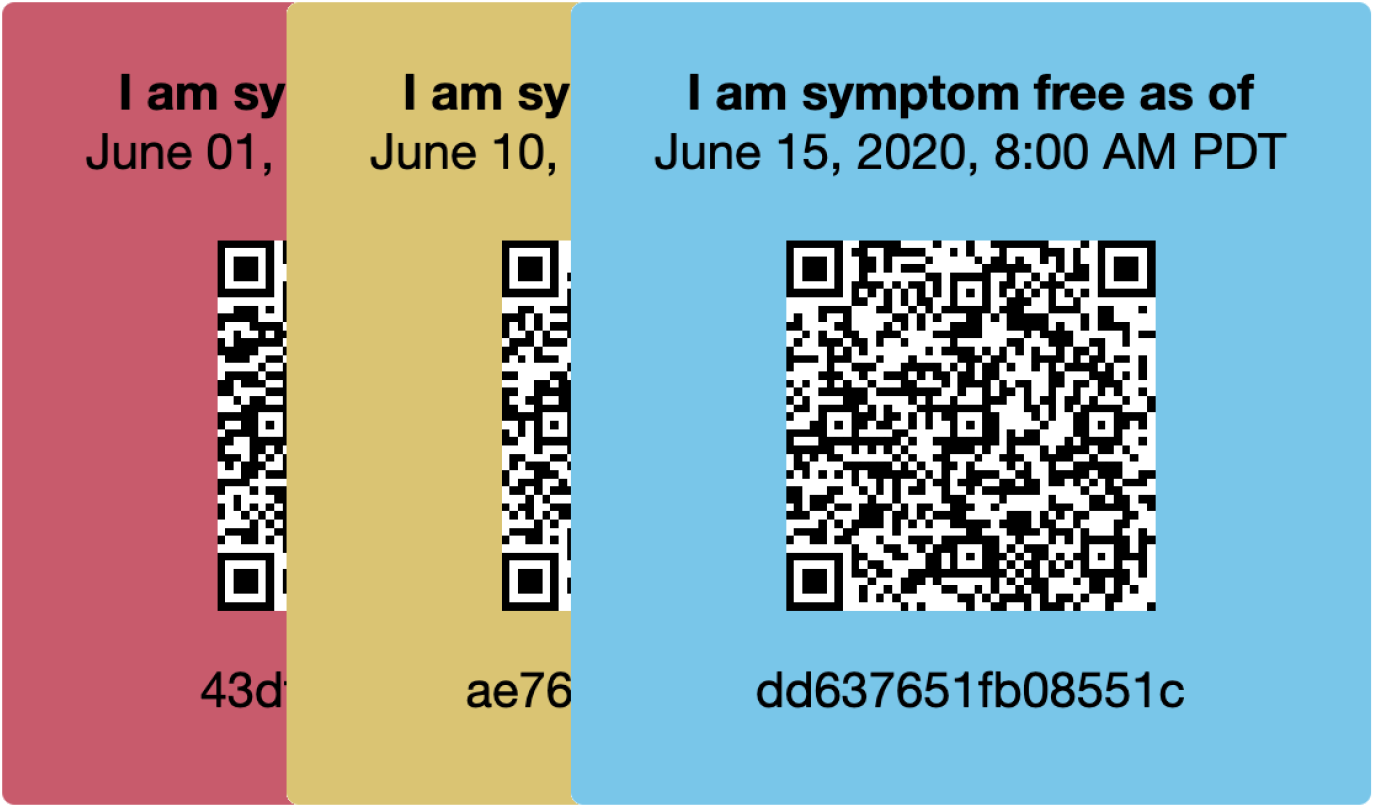
Examples of anonymous symptom free certificates.

An issue with anonymous certificates is that it is possible that a user can obtain a certificate from someone else without taking the screener and for some organizations this is a problem. Our implementation also supports named certificates where a name or other identifier is displayed on the certificate. Similar to a digital signature, the code on a named certificate corresponds both to the time the symptoms were reported and is tied to the name or identifier on the certificate. If the name is changed, then the certificate will no longer be valid. Named certificates present an additional privacy challenge in that the symptom screener must know the name to issue the right code. We address this issue by instead of using the name or identifier, we group names into equivalence classes which we call name variants. Each name or identifier is assigned a name variant which is a number between 1 and 10,000. The issued code matches the specific number. If someone modifies the name or identifier on the certificate, the number corresponding to the name will have a 99.99% chance of being different from the number of the original name which will make the certificate invalid. This prevents individuals from sharing symptom free certificates and ensures that they themselves completed the screening, further increasing the security of the system. To preserve privacy, the screener only obtains the information of the name variant or which number a specific name or identifier corresponds to and not the actual name or identifier which is all that is necessary to issue the correct code. If there are many individuals who are using the system, there will be many using it in each hour with identifiers that map to the same name variant which will provide privacy. The verification system takes as input the code and the name and will verify that the code matches the name. The verification system displays the name on the certificate if it’s valid. An example of a named certificate is shown in Figure 2 and can also be validated using the QR code.

**Figure 2.**
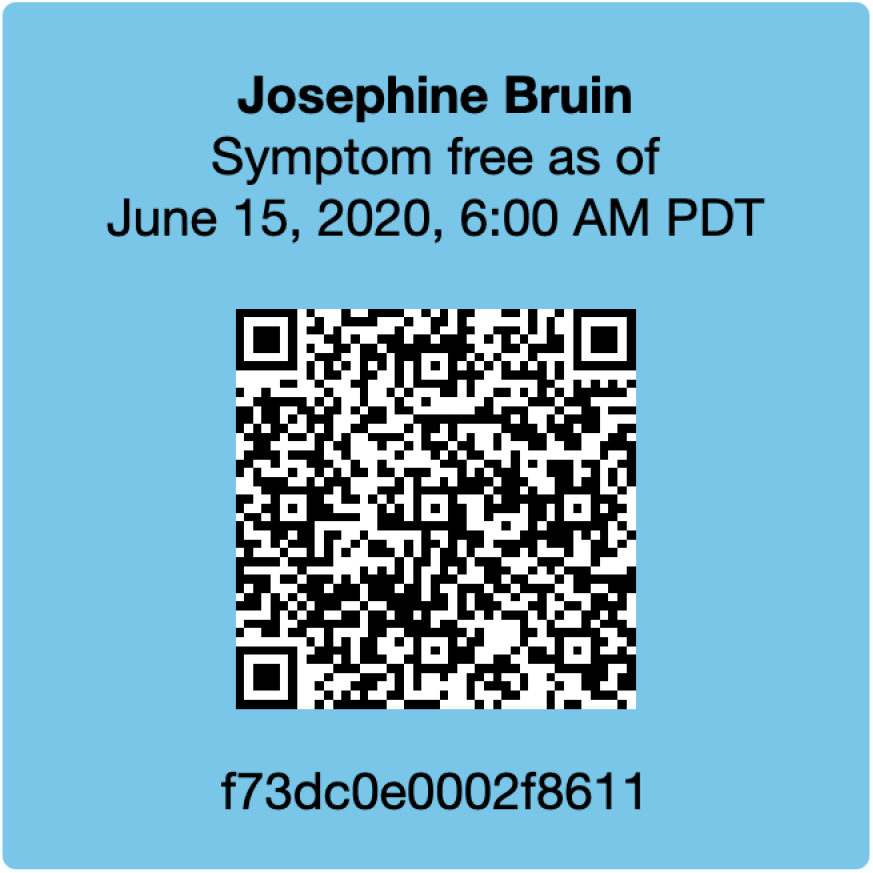
Example of named symptom free certificate.

We note that any organization can implement their own validation system which can integrate into their own internal systems. If an organization implements their own validation system, then the names or identifiers on the certificates would only be shared internally.

## CASS Cryptographic System

### Standard Digital Signatures

One key property that CASS aims to achieve is a separation between the organization and the third party screener that receives symptom information from users. Namely, the organization wants to have reasonable confidence that each employee has completed the screener, but should not be privy to the private health information that is revealed by the employee to the screener. In order to do this, we need a way for the screener to provide a certificate to the user that a survey has been completed, that the organization can check without needing to contact the screener. Cryptography offers a well-known solution to this problem, namely, digital signatures.

Digital signatures are defined by a signing and a verification algorithm. In the classic scenario, there are two parties--a signer, Sam, and a verifier, Vivian. Sam wants to send Vivian a message AND wants to guarantee the authenticity of his message where authenticity means guaranteeing the message indeed originated from Sam and was not tampered with in transit. Sam guarantees authenticity by producing a digital signature on the message that only he could reasonably produce. In our scenario, Sam has a public and private key pair. Sam runs the signing algorithm on said data or message with his private key to produce a digital signature for that piece of data or message. He then sends the signature along with the message to Vivian. Since Sam’s public key is accessible to anyone, if Vivian receives the purported message and signature from Sam, Vivian can run a verification algorithm that takes as input the message, Sam’s digital signature, and Sam’s public key. If the message from Sam is untampered and was signed with Sam’s private key, then the verification algorithm will approve the message as Sam’s. A secure digital signature scheme guarantees that with high probability, no reasonably computationally powerful malicious individual can forge a signature on some message when the malicious individual has no access to the private key of the individual they impersonate.

In our setting, the survey system generates a public-private key pair. Then, digital signatures can be signed on a completion code by the survey system when an individual completes the survey. The organization can then verify that the completion code was indeed signed by the survey system. A secure digital signature scheme guarantees that no reasonably computationally powerful malicious individual can forge a signature on a completion code.

However, as we now discuss, because usability is of paramount importance to CASS, we cannot use digital signatures as they exist today. State-of-the-art digital signature schemes require certificates to be of size at least 154 bits, which if expressed in hexadecimal, would be 39 symbols long (Boneh, Lynn, and Shacham 2004). Because users must present certificates to their organizations daily, allowing them to be embedded in URLs or even having them printed on paper makes them more practical and more likely to be used. Since the public health benefits of such a system directly depend on how compliant employees are and this it is critical that these certificates be as short as possible. In particular, we impose on ourselves a limit of at most 16 hexadecimal characters, so that even if a user must type in this certificate by hand, it would be similar to typing a credit card number (16 decimal digits). To accomplish this, we take an alternative route, which achieves a moderate level of security, as detailed below, while still allowing for very short certificates.

### Relaxed Digital Signatures

We provide moderate cryptographic security to encourage participants to take the survey. Since taking the survey is simple to do, we only provide security appropriate to the difficulty of behaving honestly. Though we could have used digital signatures or stronger security notions, doing so would have increased the size of the completion code, the runtime, and the complexity of the system. We believe that our simplicity, usability, and efficiency versus security tradeoff is appropriate since the survey takes only a few minutes to complete.

The CASS completion code generator is a software that is hosted by the third party screener. To generate completion codes, CASS first generates a secret random string r of length 16 bytes. Let K be the number of name variants supported by the system. A value N G [0, K-1] is calculated from a user’s name by computing the SHA-256 hash and reducing this value modulo K. For an anonymous name code, the value N is set to 0. Given N, and at day D and hour T that the survey is completed, CASS computes the following value A, called a “raw code”, by computing the SHA-256 hash of these values concatenated:

> A = SHA-256(0, r, T, D, N)

The output of SHA-256 is a string of hexadecimal values. The actual completion code, B, given to the user will be prefixes of the raw code. Namely, B is the first 16 characters of A. Thus, each code will look something like this: 3239c5bd01d861b6.

Without knowing r, the probability of successfully guessing a valid receipt is roughly 2^4^*^16^ to 1, or less than one in 64 trillion. Furthermore, every day, CASS also chooses, for each hour and each name code, a random integer salt between 0 and 10,000 inclusive. It then computes the following verification code, C, by prepending the salt and hashing B:

> C = SHA-256(salt, B)

Note that only 24*K values of verification codes (one for every name code) must be computed each day by CASS. There is a public website that will hold all the verification codes computed each day, which will be made public at the start of each day.

The CASS completion code validator is a software that can be executed on a website, or using desktop software. Every day, the software will obtain the list of all verification codes from the public website. When an organization receives a potential code from a user, they can enter the receipt into the software. Given this receipt, E, the software will compute potential verification codes, F, by hashing with each possible salt value:

> For i = 1 to 10,000:
>
> F = SHA-256(i, E)
>
> If F in verification codes, return T, D, N.
>
> Return error.

Each potential verification code is checked against the verification codes for each hour stored in the software. If the potential verification code matches, the software returns the day, hour, and name variant for which the code matched. If the potential code does not match any of the verification codes, then an error is displayed, and the offending code is displayed as being invalid.

### Threat Model

We again emphasize that we do not guarantee computational security against forging or cheating behavior. Our scheme makes a reasonable tradeoff for usability compared to digital signatures. Nevertheless, the CASS completion code offers some computational deterrence to a would-be forger or survey completion cheater.

Consider the following security analysis on the completion codes. Suppose an adversary wanted to gain a valid completion code without taking the survey. Using only the public verification codes, it is computationally infeasible to generate a valid completion code for a chosen name, day, and time stamp. If we model SHA-256 as a random function, then the probability of an adversary generating such a completion code is less than 2’77. In particular, this probability is computed by observing that an adversary would have to guess the random 16-byte string (probability of 2’^64^) and a salt value ranging in the interval of integers between 0 and 10,000 (probability of 2’^13^). In other words, the expected number of guesses a user would have to make is 2^77^.

Adversaries may have friends who have completed the survey and are willing to share their completion codes with the adversary. In the variant where we do not include the individual’s name, then there is nothing to stop the adversary from using one of their friend’s codes. However, in the named variant, then if we model SHA-256 as a random function, the probability that the code of any given friend is also usable by the adversary is less than 1/K, where K is the specfiable number of name classes/variants being used. In particular, in this implementation, K = 10,000. We note that a friend could take the survey under the adversary’s name, but think that the incentive for this is low.

The completion codes cannot leak any information about the survey takers apart from the hour in which they took the survey and their name (if we use the named variant of the completion code). This is because the completion code is computed independently of all other personal information.

## Discussion

We present an approach for COVID-19 symptom screening which preserves privacy enabled by cryptography. In our approach, users provide their symptom information to a third party screener and if they report no symptoms, they are issued a symptom free certificate. We developed a novel relaxed digital signature scheme to provide a code on the certificate to certify its authenticity. The scheme enables codes that are much shorter than standard digital signatures to increase the usability which will increase the compliance by users resulting in a greater positive effect on public health.

Employee symptom screening in the workplace is being mandated by many public health authorities to increase the safety of the workplace and help control infections. Privacy issues complicate the deployment of symptom screening even for employees. However, employees are not the only individuals in the workplace and for non-employees, the privacy issues are even more substantial. Since our approach does preserve privacy, our system can be easily extended to all individuals who are present in a workplace.

Since our approach preserves privacy, it can be used in many more scenarios where privacy issues are even more sensitive. One example is that our approach can provide symptom screening for retail customers or restaurant patrons. For this to be enabled, a sign outside the business would ask customers to go to the screener website to obtain their certificate which they can show upon entry. Another example is that the approach be used to symptom screen children at both daycares and schools. In this application, parents would fill out the symptom screener on behalf of their child and then show the certificate to the school or daycare when they drop their child off. These types of applications are substantially facilitated using an anonymous symptom screener as enabled by our technology.

## Data Availability

No applicable data.

## Acknowledgements

Research of A.K., P.L., and A.S. supported in part from DARPA SAFEWARE and SIEVE awards, NTT Research, NSF Frontier Award 1413955, BSF grant 2012378, a Xerox Faculty Research Award, a Google Faculty Research Award, an equipment grant from Intel, and an Okawa Foundation Research Grant. This material is based upon work supported by the Defense Advanced Research Projects Agency through Award HR00112020024 and the ARL under Contract W911NF-15-C-0205. The views expressed are those of the authors and do not reflect the official policy or position of the Department of Defense, the National Science Foundation, NTT Research, or the U.S. Government. B.J. was supported by the National Science Foundation Graduate Research Fellowship Program under Grant No. DGE-1650604.

